# Clinical and Molecular Data to Predict Flares in DMARD optimization in Rheumatoid Arthritis: A Randomised, Controlled, Open-label, Non-inferiority Trial

**DOI:** 10.1101/2025.09.30.25336957

**Authors:** Francisco J. Blanco, Laura Galindo, Belen Acasuso, Vanesa Balboa-Barreiro, Juan D Cañete, Benjamin Fernández-Gutiérrez, Isidoro González-Álvaro, José Luis Pablos Álvarez, Carmen Bejerano-Herrería Md, Maite Silva-Díaz, Ignacio Rego-Perez, Lucia Lourido, Cristina Ruiz-Romero, Miren Uriarte-Ecenarro, Rosario García-Vicuña, Andrea Cuervo, Julio Ramírez, Raquel Celis, Luis Rodríguez-Rodríguez, Lydia Abasolo Alcázar, Dalifer Freites Nuñez, Maria Martín-López, Francisco J. Toro-Santos, Natividad Oreiro

**Affiliations:** Department of Rheumatology, Rheumatology Research Group (GIR) Biomedical Research Institute (INIBIC), Complexo Hospitalario Universitario de A Coruña (CHUAC), Sergas, A Coruña, Spain; Department of Rheumatology, Arthritis Unit, Hospital Clinic Barcelona IDIBAPS, Barcelona, Spain; Department of Rheumatology and Health Research Institute (IdISSC), Hospital Clínico San Carlos, Madrid, Spain; Department of Rheumatology, Hospital Universitario La Princesa, IIS, Madrid Spain; Department of Rheumatology, Instituto de Investigación Hospital 12 de Octubre, Complutense University of Madrid, Madrid, Spain

**Keywords:** Rheumatoid arthritis, bDMARDs, flares, sustained remission, predictive models, personalized medicine

## Abstract

**Objectives:** Optimization of biological DMARDs (bDMARDs) in patients with rheumatoid arthritis (RA) may be feasible in those who have maintained remission for at least six months. However, this approach carries the risk of disease flare, underscoring the need for reliable predictors to guide clinical decisions.

**Methods:** The OPTIBIO trial (EudraCT 2012-004482-40) was a phase IV, randomized, open-label, non-inferiority study conducted in five hospitals in Spain. RA patients in sustained remission on stable bDMARD therapy were randomized 1:1 to standard care or dose reduction. The primary outcome was to compare to proportion of joint flare at 12 months by a non-inferiority analysis analyzed by the intention-to-treat principle and identify predictors for flare and sustained remission.

**Results:** 195 patients were randomized: 99 to the control group and 96 to the optimization group. Thirty-nine flares occurred (optimization: 22.7%, control: 17.2%), with a risk difference of -5.5% (95% CI: -16.8% to 5.7%; P = 0.33). Two predictive models were developed: one for flares (AUC: 0.84) including 3v-DAS28-CRP, VAS pain, erosions, systolic blood pressure, and hemoglobin, and another for sustained remission (AUC: 0.77) including 3v-DAS28-CRP, age, and rheumatoid factor. Adding molecular biomarkers improved AUCs to 0.91 and 0.88, respectively. No significant differences in adverse events were observed.

**Conclusion:** bDMARD dose optimization was non-inferior to standard therapy on the flare rate but demonstrated similar safety. Predictive models for remission and flares were developed, which may help select patients to ensure safe implementation of this strategy, highlighting the need for personalized treatment.

**Key messages:**

- bDMARD dose optimization was non-inferior to standard therapy on the flare
- Clinical models classified patients in terms of disease severity
- Incorporation of molecular markers to clinical models improved their prediction power

## Introduction

The RA treatment aims to achieve remission or maintain low disease activity (LDA) to prevent irreversible joint damage, reduce disability, and enhance patients’ quality of life.^1^ The American College of Rheumatology (ACR) and the European Alliance of Associations for Rheumatology (EULAR) recommend a treat-to-target (T2T) strategy, which emphasizes regular monitoring of disease activity and therapy adjustments to sustain optimal disease control.^2,3^

The introduction of bDMARD and Janus kinase inhibitors (JAKi) has significantly improved treatment outcomes, enabling more patients to maintain disease remission. This has led to growing interest in the potential for tapering biologic therapies to reduce medication exposure and associated risks while preserving disease control. Observational studies have examined the feasibility of dose reduction, reporting variable adherence to tapered regimens and differing impacts on disease stability.^4,5^

The ACR and EULAR guidelines provide recommendations on bDMARD dose reduction in RA^2,3^. The American guideline conditionally favors continuing the dose due to low-certainty evidence,³ whereas the European guideline recommends reduction only after glucocorticoid discontinuation and at least 6 months of sustained remission.² Despite these recommendations, further data are needed to improve prediction and prevent flares.

Randomized controlled trials (RCTs) have investigated tapering strategies to determine whether reduced dosing can maintain disease control comparable to standard therapy. Findings, however, have been inconsistent across different biologics. Some RCTs reported a higher risk of flares and a reduced likelihood of maintaining remission,^6, 10^ while others found no statistically significant increase in risk ^11, 12^. Factors such as treatment type, patient characteristics, and disease history appear to influence tapering success, underscoring the need for personalized approaches.

Efforts to identify predictors of flare during dose reduction have highlighted several clinical factors linked to increased or decreased relapse risk, including anticitrullinated protein antibodies (ACPA), rheumatoid factor (RF), corticosteroid use, gender, and imaging evidence of subclinical inflammation ^13–22^. However, no single predictor has proven universally reliable, underscoring the need for a precise approach to guide tapering decisions.

Given the variability in outcomes, further research is needed to refine patient selection criteria and develop strategies that balance efficacy and safety. Identifying robust predictive markers could enable personalized tapering protocols, minimizing flare risk while optimizing long-term disease control in RA patients.

The aim of this study was to compare the proportion of joint flare at 12 months by a non-inferiority analysis and identify predictors to optimize treatment and personalize care.

## Methods (Please ensure information on ethical approval and informed consent is provided)

### Patients and Methods

The Biologic Optimization Study (OPTIBIO) was a Phase IV, randomized, controlled, open-label, non-inferiority trial conducted in patients with RA in sustained remission on biological therapy. The recruitment occurred at five hospitals in Spain. The study adhered to ethical principles in biomedical research and the current legislation in Spain, following Good Clinical Practice (ICH Topic E6, 2011), the Declaration of Helsinki, and the requirements of RD 223/2004. The Autonomous Ethics Committee of Galicia (CEIC) approved the study, and all patients provided written informed consent.

Eligible patients were required to be at least 18 years, RA diagnosis according to the ACR 1987 criteria and have been in remission with stable bDMARD therapy for at least six months prior to screening. Remission was defined using one of the following: a Disease Activity Score 28 (DAS28) ≤2·6 (including C-reactive protein [CRP], erythrocyte sedimentation rate [ESR], or those with three variables), a Simplified Disease Activity Index (SDAI) ≤3·3, or meeting ACR/EULAR Boolean criteria.

Use of conventional synthetic DMARDs (csDMARDs) was permitted, on stable doses for at least six months prior to inclusion. Oral steroids at doses ≤10 mg/day of prednisone (or equivalent) were also eligible. Those who had received intra-articular steroid injections within the past six months were excluded.

### Randomization

Patients were randomly assigned in a 1:1 ratio to either optimize their bDMARD therapy or continue with the standard dose. Randomization was centralized using a stratified allocation method based on each patient’s current treatment. Allocation was concealed until the randomization process was completed to ensure impartiality.

### Procedures

Patients in the optimization group reduced their bDMARD therapy by extending the dosing interval or by reducing the dose, while the control group continued treatment at the guideline recommended (Supplementary material 1). They were followed up with visits every 12 weeks and additional visits scheduled for flares or any events of interest.

Patients who experienced a joint flare in the optimization group returned to their baseline bDMARD dose, while those in the control group received care per current treatment guidelines. No re-optimization was attempted in patients who regained remission after treatment reintroduction.

### Outcomes

The primary outcome was to assess the flare-rate between baseline and 12 months through a non-inferiority analysis. Disease activity was evaluated at each centre by an independent, blinded rheumatologist. A joint flare was defined as an increase of DAS28 >2·6 points, SDAI >3·3 points and the loss of the ACR/EULAR Boolean remission criteria. Patients who maintained a DAS score less than 2·6, SDAI <3·3, or met ACR/EULAR Boolean remission criteria were defined as sustained remission.

Secondary endpoints were to compare flare-free survival at 18, 24, and 36 months; to develop two prediction models, one for flare and one for sustained remission; and to evaluate the accuracy of each model. Finally, potential clinical and laboratory adverse events were assessed.

### Targeted next-generation sequencing

Serum biomarkers were collected at baseline visit. The Ampliseq IAD211967_182 panel was used to sequence 22 selected genes based on extensive curation of peer-reviewed literature and disease research databases (Supplementary material 2). 195 DNA samples were analyzed (96 optimization group, 99 control). Libraries were prepared using Ion Chef and sequenced on an Ion S5XL, obtaining ∼20 million reads per chip with an average depth of 1300X.

### Analysis of sequencing data

Reads were aligned to GrCh37 using Torrent Suite. Variants were called with low stringency, filtered by depth, PHRED score, and frequency. Aggregated vcfs were filtered by MAF < 0.01. Plink converted data for logistic regression on flare/remission, including the additive effect of the allelic dose of each variant. Results were annotated using SnpEff.

### Anti-cytokine Autoantibody (ACAA) Analysis

The MILLIPLEX MAP panel was used to detect and quantify 15 anti-cytokine autoantibodies in serum using xMAP technology. Samples were diluted, incubated, and analyzed on a MagPix instrument. Assay precision was assessed by calculating the coefficient of variation for replicate samples withing each plate. (The complete molecular analysis is found in the Supplementary material 3-5).

### Statistical Analyses

A total sample size of 195 patients was calculated. This sample size allowed us to evaluate the non-inferiority hypothesis with a power of 80% and an alpha value of 0·05 with the assumption of a remission rate at one year of 92% in the standard therapy group, a non-inferiority margin of 10% and considering a 10% loss to follow-up.

Baseline characteristics were described by frequencies and percentages for categorical variables and mean (SD) and median (IQR) for continuous variables. The student’s T test or Mann-Whitney U test was used to compare means between treatment groups, depending on the normality analysis with the Kolmogorov-Smirnov test. The Pearson chi-square test or Fisher’s exact test was applied to assess the relationship between qualitative variables.

For the primary and safety analyses, data were analyzed using the intention-to-treat (ITT) population. The proportion of joint flares at 12 months was compared between the two groups using the Chi-square test. Flare-free survival was assessed using the Kaplan–Meier method, and hazard ratios (HRs) were estimated through a Cox proportional hazards model, using the control group as the reference. Differences between survival curves were evaluated using the log-rank test.

Secondary outcomes were analyzed by the same methodological approach used in the primary analysis for the proportion of flares and flare-free survival analyses at 18, 24 and 36 months.

Univariate and multivariate Cox proportional hazards regression analyses were conducted to evaluate the risk of joint flare, HRs and their corresponding 95% CI reported. Additionally, univariate and multivariate logistic regression analyses were performed to assess the odds of achieving sustained remission, with odds ratios (ORs) and 95% CI calculated. For the selection of the final clinical model, the variables with p<0·20 in the univariate models and those clinically relevant according to the literature were included. Manual elimination of covariates was carried out backwards until the final model was obtained. Subsequently, the complete model was obtained using clinical and molecular variables. Finally, an internal validation of the models was performed using a bootstrap method (500 replicates) to calculate the corrected AUC, with the aim of evaluating the predictive accuracy of the models at 12 months. An AUC exceeding 0·75 was interpreted as indicative of good predictive performance. The optimization group was selected to perform the prediction models. All statistical analyses were conducted using SPSS v28 and R v4.3.1 software. Statistical significance level of p < 0·05 was considered. This trial was registered in EudraCT with identification number: 2012-004482-40 (www.clinicaltrialsregister.eu).

This manuscript used ChatGPT to improve the clarity and grammar of the language.

## Results

Two hundred and forty one patients were evaluated for eligibility. Forty-six were excluded before randomization: 17 presented a flare, 18 did not meet all inclusion criteria, five for patient decision, five for medical decision and one for missing data. A total of 195 patients were enrolled and randomly assigned: 99 to control group and 96 to optimization group. During the first year of follow-up, four patients in optimization group and three in the control group were lost (Figure 1).

**Figure 1:**
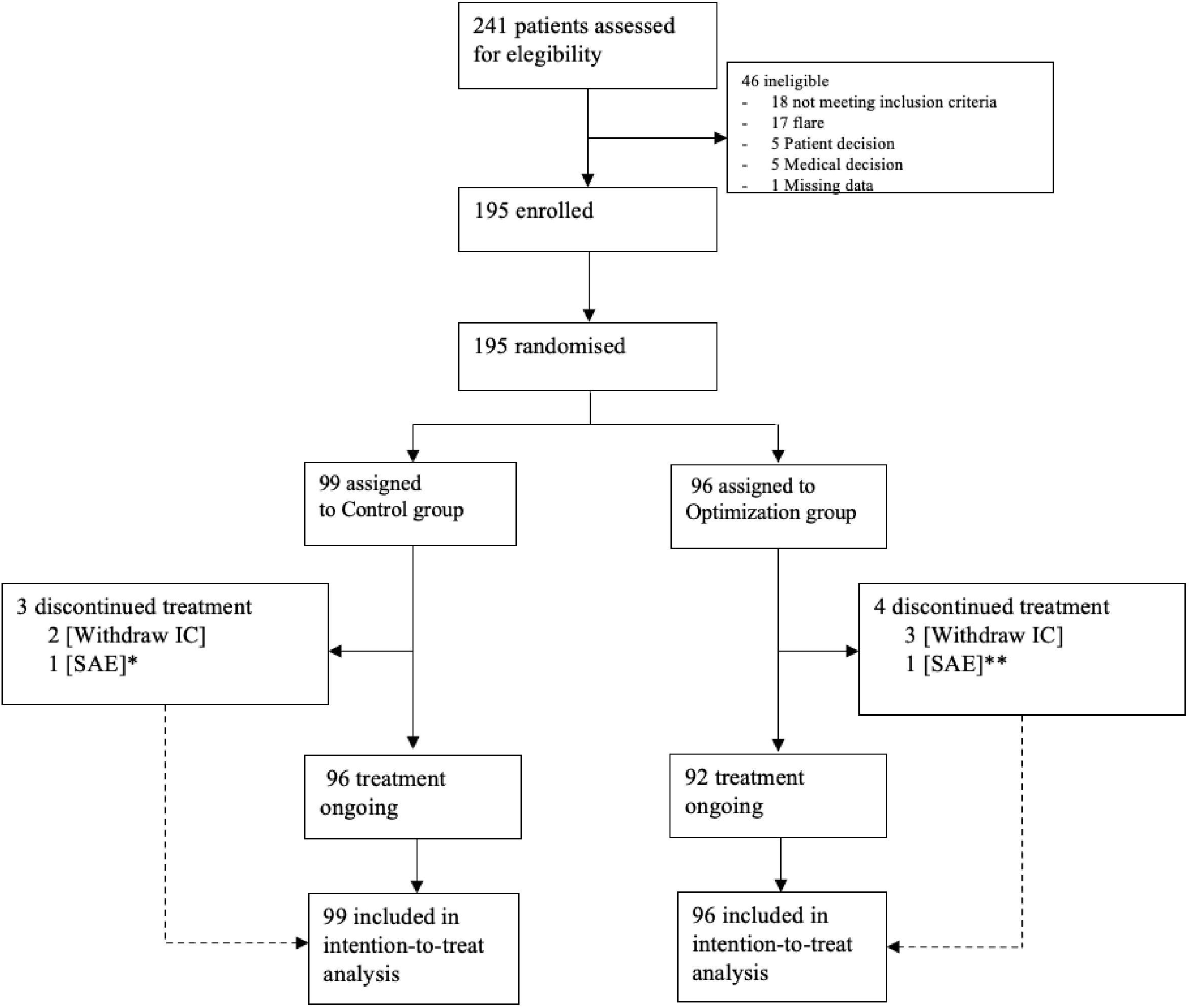
Flowchart of patients. IC = informed consent * Cutaneous Melanoma ** Space Occupying Lesion of the Brain

Baseline characteristics were comparable between groups (Table 1). The mean age of participants was 63 years (SD 12.78), and 83% were women. The mean remission duration was 22.19 months (SD 16.07) for the control group and 20.82 months (SD 14.55) for the optimization group.

**Table 1:**
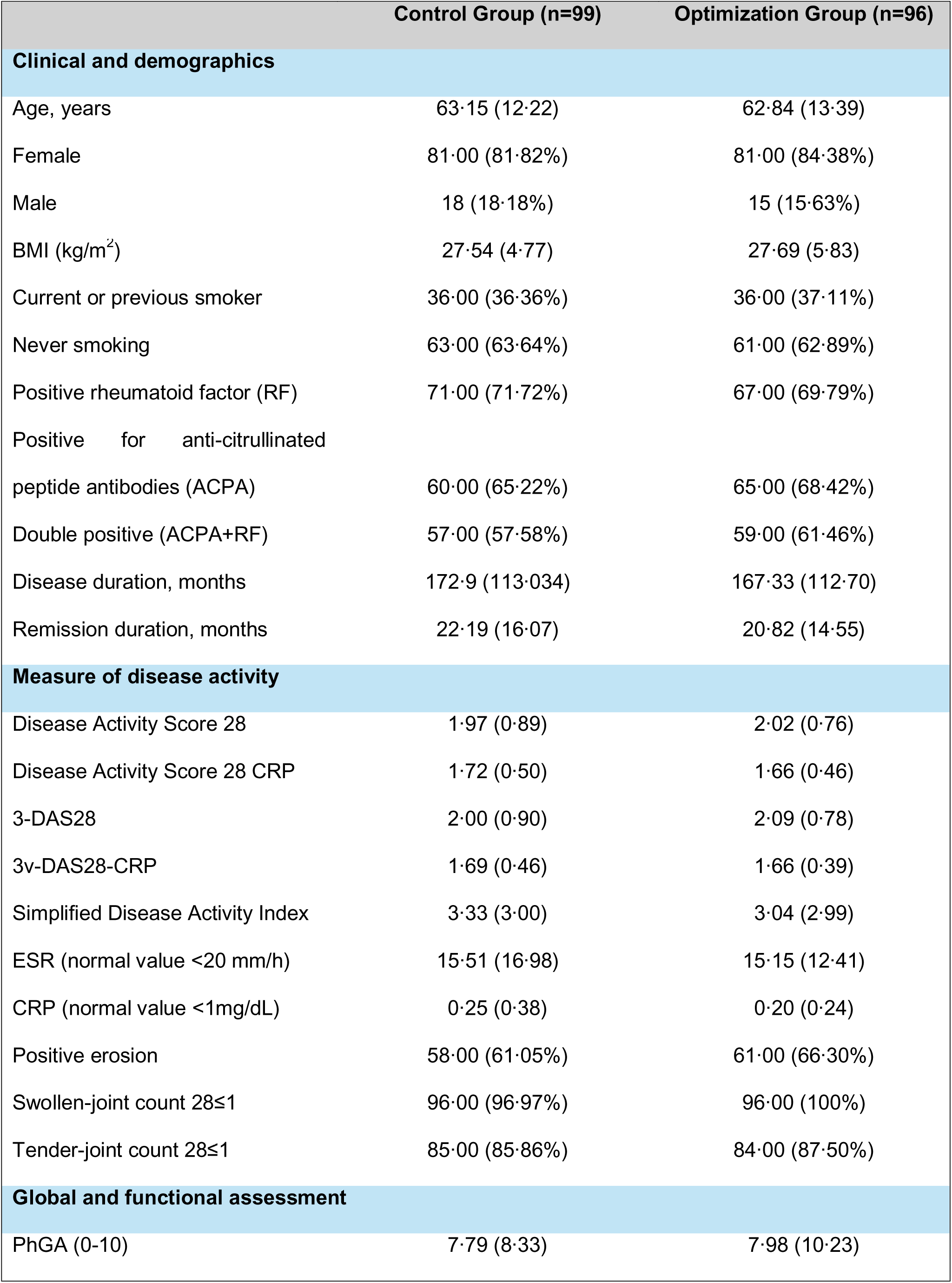
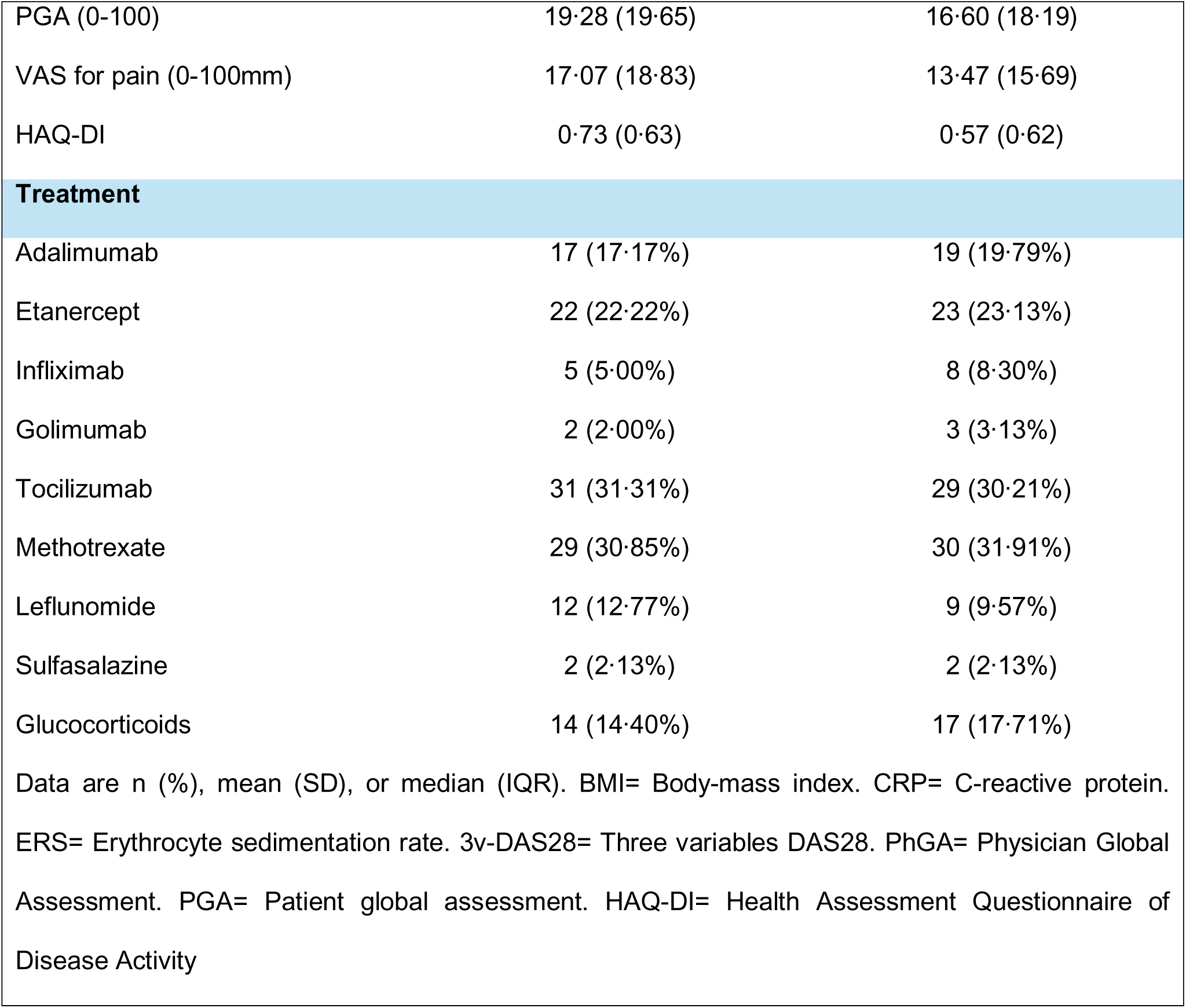
Baseline characteristics of the study population.

At 12 months, 39 flares were recorded: 17 (17.2%) in the control group and 22 (22.7%) in the optimization group, yielding a risk difference of –5.5% (–16.8 to 5.7; p = 0.33). However, the lower limit of the 95% confidence interval exceeded the non-inferiority margin of –10%. Thus, non-inferiority could not be demonstrated at 12 months (Figure 2a).

**Figure 2a:**
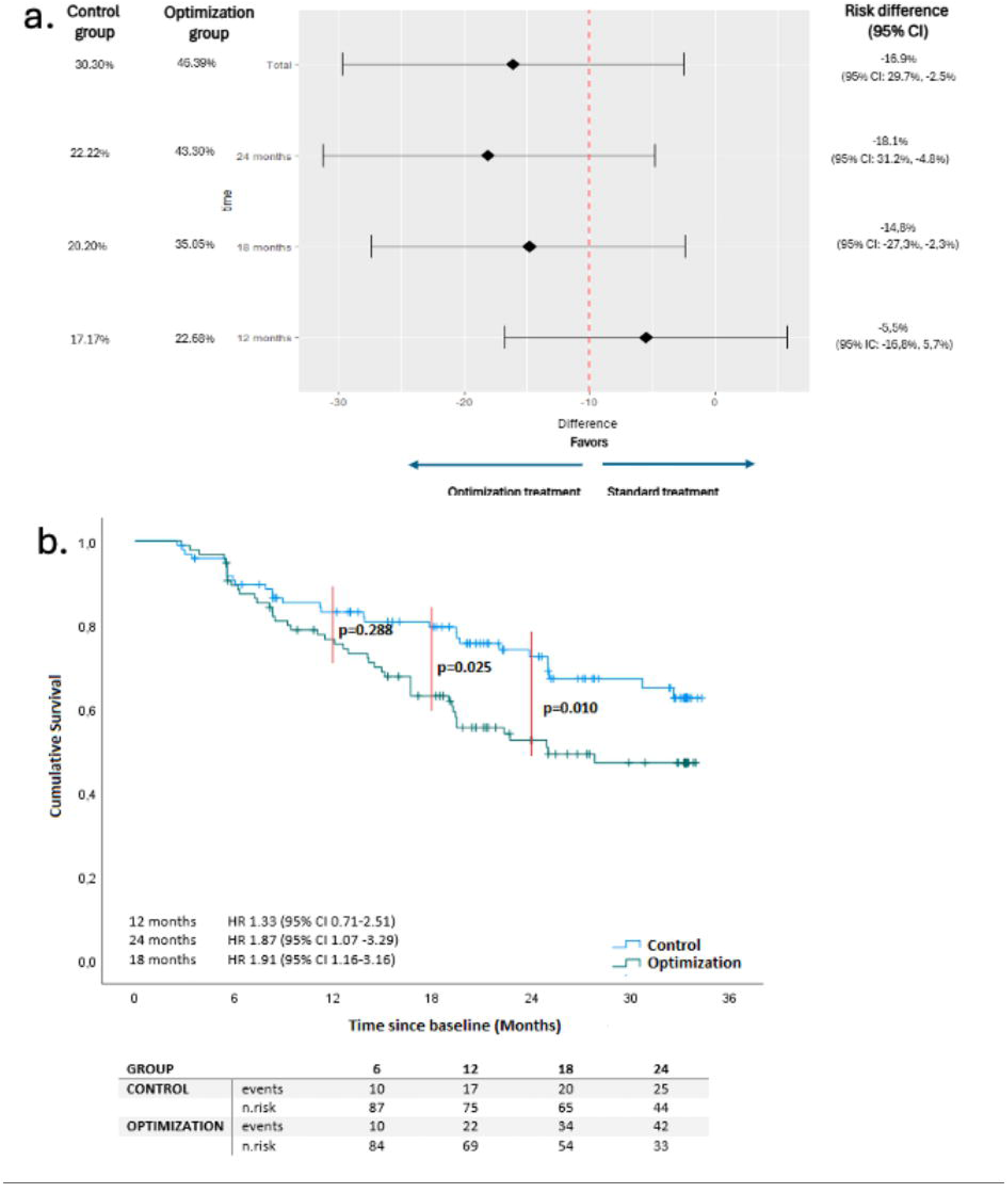
NON-INFERIORITY ANALYSIS OF THE COMPARISON OF FLARE RATE (PRIMARY OBJECTIVE) AND ADDITIONAL ANALYSES. The primary endpoint was the proportion of relapses in each group using non-inferiority analysis. The dotted vertical red line represents the non-inferiority margin (10%). At 12 months, the 95% confidence interval exceeded the established lower limit; this result was maintained throughout the study. Figure 2b**: FLARE-FREE SURVIVAL CURVE THROUGHOUT FOLLOW-UP.** The analysis was done in the intention to treat population. Red lines indicate the analyses at each time point. No significant difference was found between the survival of both groups at one year of follow-up. Flare-free survival was higher in the control group and was statistically significant from month 18 of follow-up. The risk of flare was analyzed by a Cox Proportional Hazard Regression with 95% CI represents the risk for the optimization group compared to the control group.

Analysis of flare-free survival showed a HR=1.33 (0.71–2.51) at 12 months, which did not reach statistical significance. However, at 18 and 24 months the flare-free survival was statistical significant (Figure 2b).

In Univariate Cox regression, older age, higher baseline disease activity scores (DAS28-CRP, 3v-DAS28-CRP, and SDAI), tender joint counts, elevated CRP levels, higher Visual Analog Scale (VAS) pain, and greater disability were significantly associated with a risk of flares. On a molecular level, joint flares were associated with seven single nucleotide polymorphisms (SNPs) in genes related to disease activity or treatment response (Supplementary material 6-7).

In the clinical model, higher disease activity (3v-DAS28-CRP) emerged as the strongest predictor (HR=14.53 (2.82–74.73)). Elevated hemoglobin levels and the presence of erosions were also associated with increased risk (HR=1.77 (1.09–2.89) and, HR=7.28 (1.58–33.55), respectively). In addition, both systolic blood pressure (SBP) and VAS pain contributed with modest effects (HR=1.04 (1.00–1.07) and HR=1.04 (1.01–1.07), respectively (Table 2).

**Table 2:** Joint flare prediction models.

|  | HR | (IC, 95%) | p value |
| --- | --- | --- | --- |
| **Clinical model** |  |  |  |
| 3v-DAS28-CRP | 14.53 | 2.82-74.73 | 0.001 |
| Hb (g/dL) | 1.77 | 1.09-2.89 | 0.020 |
| Erosion | 7.28 | 1.58-33.55 | 0.010 |
| SBP (mm/Hg) | 1.04 | 1.007-1.07 | 0.016 |
| VAS pain | 1.04 | 1.01-1.07 | 0.007 |
| **Complete model** |  |  |  |
| 3v-DAS28-CRP | 7.28 | 1.43-36.95 | 0.016 |
| Hb (g/dL) | 1.52 | 0.88-2.60 | 0.125 |
| SBP (mm/Hg) | 1.02 | 1.001-1.05 | 0.040 |
| Erosion | 8.15 | 1.58-42.02 | 0.012 |
| VAS pain | 1.06 | 1.02-1.10 | <0.001 |
| rs5746065 (C/A) | 0.31 | 0.09-1.004 | 0.050 |
| rs9594987 (T/C) | 0.15 | 0.04-0.57 | 0.005 |
| Anti-IFNγ | 1.0005 | 1.00004-1.0009 | 0.031 |
| 3v-DAS28= Three variables DAS28, Hb = Hemoglobin, SBP = Systolic Blood Pressure, VAS = Visual Analogue Scale |  |  |  |

After adding molecular variables, 3v-DAS28-CRP and erosions remained as strong predictors (HR=7.28 (1.43–36.95) and HR=8.15 (1.58–42.02)). Pain intensity retained significance (HR= 1.06 (1.02–1.10)), while SBP showed a smaller effect (HR=1.02 (1.00–1.05)). Hemoglobin, however, was no longer significant (HR=1.52 (0.88–2.60)). Among molecular variables, rs5746065 (C/A) in TNFRSF1B gene showed a borderline protective effect (HR=0.31 (0.09–1.00)), whereas rs9594987 (T/C) in ENOX gene was significantly protective (HR=0.15 (0.04–0.57)). Anti-IFNγ levels were also linked to increased risk (HR=1.00 (1.00–1.00)) (Table 2). The model demonstrated good discriminative performance at 12 months, with a concordance C-index of 0.884, and was highly significant overall χ² = 44.18, 8 df; p = 5×10⁻⁷.

The most accurate clinical model yielded an AUC=0.84 (0.71–0.97). The inclusion of molecular biomarkers improved the model’s discriminatory capacity, increasing the AUC value to 0.91 (0.82– 0.99). This model demonstrated a sensitivity of 75% and specificity of 93%, with negative and positive predictive values (NPV/PPV) of 93% and 76%, respectively (Figure 3a-3b).

**Figure 3.**
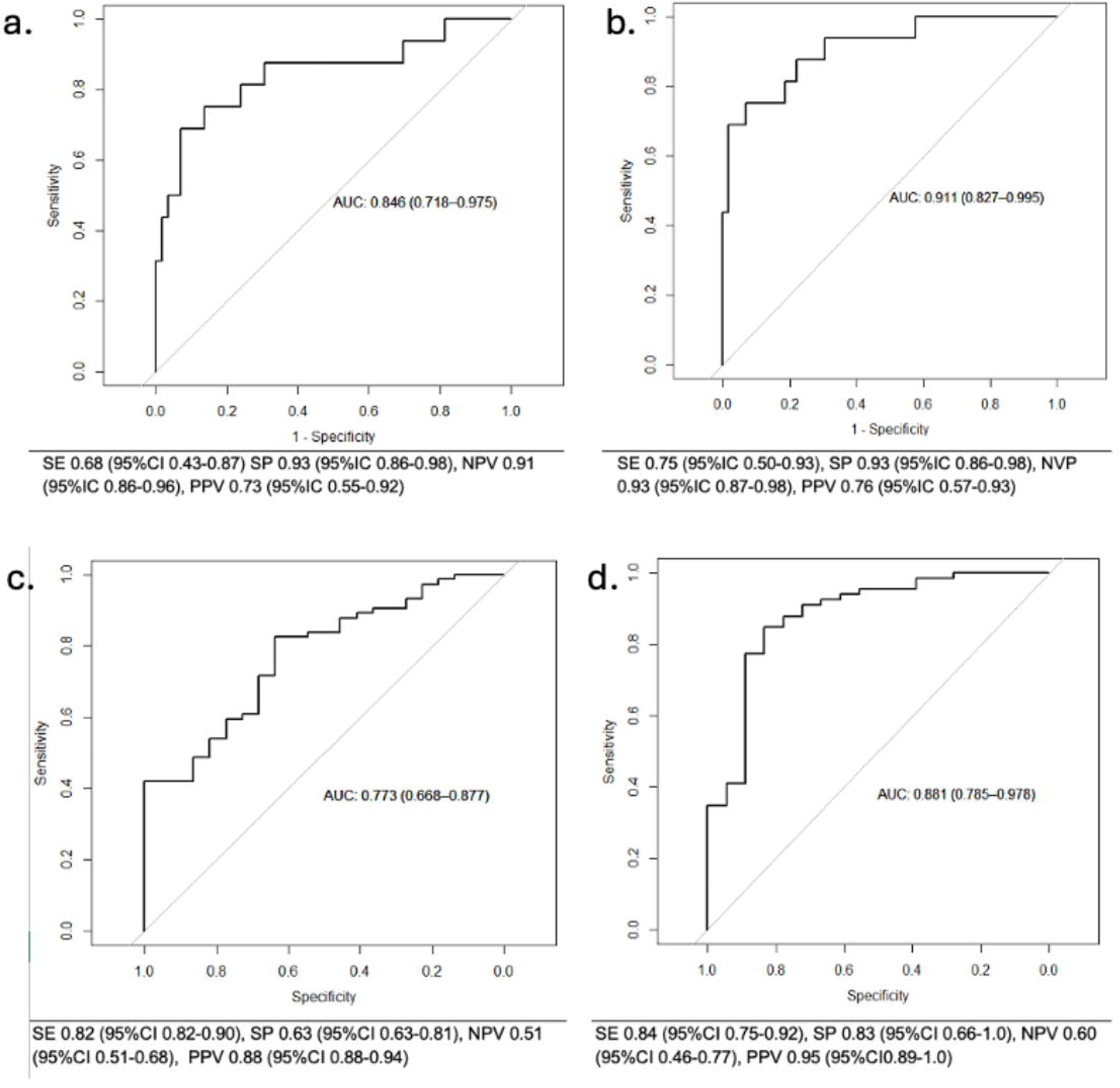
ROC CURVE OF THE CLINICAL AND COMPLETE SUSTAINED REMISSION AND FLARE PREDICTION MODELS. Figure 3a shows an area under the curve (AUC=0.77 (0.66-0.87)). Sensitivity (SE) of 82%, a specificity (SP) of 63%, a positive predictive value (PPV) of 88% and a negative predictive value (NPV) of 52%. Figure 3b. AUC=0.88 (0.78-0.97) SE of 84%, a SP of 83%, a PPV of 95% and a NPV of 60%. The curve represents the relationship between sensitivity and 1-specificity, evaluating the discriminative performance of the model. Figure 3c shows an AUC=0.84 (0.71-0.97). SE of 68%, SP of 93%, PPV of 73% and a NPV of 91%. Figure 3d. AUC=0.91 (0.82-0.99). SE of 75%, a SP of 50%, a PPV of 76% and a NPV of 93%. The curve represents the relationship between sensitivity and 1-specificity, evaluating the discriminative performance of the model. All articles published in Oxford University Press journals must now include alt text (alternative text) alongside their main figures. This is to ensure that individuals with visual impairments or those using screen readers can comprehend the content and context of your figures. Alt text is applicable to all images, figures, illustrations, and photographs. Alt text is only accessible via e-reader and so it won’t appear as part of the typeset article. Could you please provide alt text for all figures included in your main manuscript? Please see this <u>OUP guide</u> for more information and examples.

For sustained remission, univariate analysis showed that age was negatively correlated with remission. In contrast, a higher heart rate was positively associated with it. Pain intensity and the number of painful joints were also negatively correlated. Similarly, greater disease activity (3v-DAS28-PCR) and higher disability scores (HAQ-DI) reduced the likelihood of maintain remission. Additionally, seven SNPs; were significantly associated with the probability of remission (Supplementary material 8-9).

In the multivariate logistic regression model, higher DAS28, remained significantly associated with a reduced likelihood of remission. Specifically, each unit increase in DAS28 corresponded to a 90% decrease in the odds of remission. Age also showed a significant negative effect, with each additional year decreasing the odds of remission by 7%. In contrast, the presence of RF was positively associated with remission, increasing the odds by more than threefold (Table 3).

**Table 3.** Sustained remission prediction models in the optimization group.

|  | OR | (IC 95%) | p value |
| --- | --- | --- | --- |
| **Clinical model** |  |  |  |
| Age (years) | 0.93 | 0.88-0.97 | 0.007 |
| Positive rheumatoid factor | 3.47 | 1.04-12.34 | 0.044 |
| 3v-Disease Activity Score 28 CRP | 0.10 | 0.01-0.45 | 0.005 |
| Age (years) | 0.93 | 0.88-0.97 | 0.007 |
| **Complete model** |  |  |  |
| Age (years) | 0.93 | 0.88-0.97 | 0.007 |
| Positive rheumatoid factor | 3.47 | 1.04-12.34 | 0.044 |
| 3v-Disease Activity Score 28 CRP | 0.10 | 0.01-0.45 | 0.005 |
| rs5746065 (C/A) | 5.77 | 1.30-29.38 | 0.024 |
| rs6555900 (G/C) | 2.30 | 0.99-5.83 | 0.059 |
| rs1560011 (G/A) | 3.27 | 1.09-11.73 | 0.046 |
| CRP= C reactive protein |  |  |  |

The complete model to predict remission at 12 months incorporating both clinical and molecular variables confirmed the significant associations observed in the initial analysis. 3v-DAS28-PCR remained a strong predictor (OR=0.10 (0.01–0.67)). RF continued to show a positive association, increasing the odds of remission by fourfold (OR=4.86 (0.98-28.67)), while age retained its negative effect, with each additional year reducing the odds by 8% (OR=0.92 (0.86–0.97)). Three SNPs were also significantly associated with sustained remission. The rs5746065 (C/A) in TNFRSF1B gene had a strong positive association, with an OR=5.77 (1.30–29.38), indicating an almost six-fold increase in the likelihood of remission. Similarly, rs6555900 (G/C) in KCNIP gene and rs1560011 (G/A) in CLEC2D gene, with odds of 2.30 (0.99-5.83) and 3.27 (1.09-11.73) respectively (Table 3). The best clinical model showed good discrimination with an AUC=0.77 (0.66–0.87). Adding molecular biomarkers improved performance, increasing the AUC to 0.88 (0.78–0.97), with 84% sensitivity, 83% specificity, and higher predictive values (Figure 3c–3d).

Regarding safety outcomes, no significant differences were observed between the reduced-dose and standard therapy groups (Supplementary material 10).

## Discussion

The OPTIBIO study did not demonstrate non-inferiority of bDMARD dose reduction compared with continued therapy at 12 months, and flare rates were significantly higher at 18 months in the optimization group. Flare-free survival was also superior with standard therapy, suggesting that long-term dose reduction may compromise clinical stability. Safety outcomes were comparable between groups.

The 12-month flare rate in our study was 22%, a figure that aligns with previously reported values in the literature. Prior studies have described flare rates following TNFi dose reduction ranging from 12% to 63%, depending on differences in dose tapering strategies, patient characteristics, and definitions of flares used across studies⁶,²². Our findings are consistent with trials such as ARCTIC REWIND, which also failed to show non-inferiority with dose reduction, though reported a higher flare rate (63%), likely due to a more aggressive tapering protocol⁶.

Conversely, some studies suggest that dose reduction may not increase flare risk under specific conditions, indicating a potential for non-inferiority when strict eligibility and monitoring protocols are applied¹¹,¹². However, such comparisons remain challenging due to heterogeneity in study design, inclusion criteria, and definitions of remission and flare. Importantly, the non-inferiority margin used in our trial was 10%, a more stringent threshold compared to the 20% margin commonly employed in other studies ⁶. This conservative approach enhances the clinical relevance and methodological rigor of our findings by setting a higher bar for demonstrating this effect, which is very important to define any predictive tool.

A meta-analysis by Uhrenholt et al., further supported by Henaux et al., concluded that bDMARD or tsDMARD dose reduction is associated with an increased risk of flare or loss of remission²³,²⁴. Although these associations did not always reach statistical significance due to heterogeneity, another systematic review corroborated these findings and underscored the importance of careful patient selection in implementing dose optimization strategies ^25^.

To better predict flare risk after dose reduction, we developed two models: a clinical model and a complete model (incorporating clinical and molecular data). The clinical model identified traditional predictors such as higher disease activity (3v-DAS28-PCR), greater disability (HAQ-DI) and structural damage, all associated with increased flare risk, even in patients deemed to be in remission. This supports prior literature where markers of residual inflammation, even at subclinical levels, increase the likelihood of relapse¹⁶,¹⁸,¹⁹,²². Interestingly, higher hemoglobin levels and higher SBP were also associated with an increased risk of flare in our study. This finding regarding hemoglobin appears to contrast with previous reports, where lower hemoglobin levels have been associated with higher disease activity, reflecting the well-established link between chronic inflammation and anemia. Regarding SBP, our findings are supported by evidence linking chronic systemic inflammation with increased cardiovascular risk, including the development of hypertension^26^.

The complete model added two SNPs and anti-IFN-γ levels, improving its predictive capacity. Elevated anti-IFN-γ levels emerged as a potential biomarker for flares, suggesting new avenues for biomarker discovery ^14,30^.

One of the identified SNPs, rs9594987 in the ENOX1 gene, has been previously associated with RA severity and potential response to tocilizumab, although the evidence remains conflicting²⁷. The inclusion of such genetic markers may improve future efforts in personalized medicine and flare prevention.

Parallel to the flare models, we developed two models to predict sustained remission following dose reduction. In the clinical model, younger age, RF seropositivity, and lower baseline 3v-DAS28-PCR scores were all associated with a higher likelihood of maintaining remission. These findings underscore the relevance of achieving deep remission before attempting dose tapering and support the prognostic value of serological markers in RA. Interestingly, contrary to previous studies⁹,¹³,¹⁶, variables such as disease duration, time to remission, ACPA seropositivity, and gender were not significantly associated with remission in our model. This highlights the variability and limitations of conventional clinical predictors across different cohorts.

The complete model for sustained remission incorporated three SNPs, improving the model’s accuracy. Among these, rs1560011 in the CLEC2D gene has been previously studied in relation to tocilizumab response and low disease activity²⁸. A novel finding was the rs6555900 variant in the KCNIP1 gene, which encodes a potassium channel regulator, not previously described in RA. Additionally, the rs5746065 variant in the TNFRSF1B gene, encoding the TNF-α receptor 2 (TNFR2), was associated with a double protective effect: reduced flare risk and increased likelihood of sustained remission²⁹.

These results support the potential utility of molecular data in identifying patients who may safely undergo dose tapering, particularly when conventional clinical markers are insufficient.

This study has several strengths. First, its randomized controlled trial (RCT) design enhances internal validity and supports the robustness of the primary outcome findings. Additionally, the extended follow-up period of up to 36 months allowed for the evaluation of both early and delayed flares, providing a more comprehensive view of the long-term impact of dose reduction. Another important strength is the development of predictive models for flare and sustained remission, which integrated both clinical and molecular variables, by using a stringent threshold of 10% to illustrate non-inferiority, with the objective of creating a model to predict the likelihood of flares and remission at 12 months.

Nonetheless, several limitations should be acknowledged. The open-label design may have introduced performance or expectation bias, although this risk was mitigated by employing blinded rheumatologists to assess disease activity. The lack of imaging data could have limited the detection of subclinical inflammation, which may precede clinical flares and could impact the interpretation of remission status. Finally, the genetic analyses were conducted exclusively in a Caucasian population, which may limit the generalizability of the molecular findings and highlights the need for validation in more diverse populations.

In conclusion, bDMARD dose reduction was not non-inferior to continued therapy on the flare rate, although safety profiles were comparable. The development of accurate clinical and molecular models to predict both flare risk and sustained remission represents an important step toward precision medicine in RA. These tools may guide personalized treatment decisions, allowing safe implementation of tapering strategies in appropriately selected patients.

## Data Availability

All data produced in the present study are available upon reasonable request to the authors

## Acknowledgements (if applicable)

Group authorship list (if applicable **and** all group members fulfil ICMJE criteria for authorship):

## Funding (if specific to this study)

This study has been funded by Instituto de Salud Carlos III (ISCIII) through the projects PMP15/00032 co-funded by the European Regional Development Fund ‘A way to make Europe’; funded by ISCIII, co-funded by the European Union and projects PI20/00793 and RD21/0002/0009 funded by Instituto de Salud Carlos III–European Union-NextGenerationEU. We also counted with funding from grant IN607A2021/07 from Xunta de Galicia. LG is a PhD student supported by Cátedra FSR-UDC, Universidad de A Coruña.

## Conflict of interest statement (in addition to author disclosure form)

Dr. Francisco Blanco has received grants or contracts from speaker fees from Abbvie, Bristol Myers Squibb, Roche, Servieer, Novartis, Horizon Therapeutics Ireland DAC, ITF RESEARCH PHARMA S.L.U., GSK Research, Pfizer, Sanofi-Aventis, Grunenthal, Lilly, Merck Healthcare, KgaA, LG Chem, Ltd, UCB, Janssen, Amgen, Regeeron, Alkem Labortories Ltd, Grunenthal, Sun Pharma Global FZE, Kiniksa Pharmaceuticals, GmbH, speakers fees from Medicamenta-Ecuador, Grunenthal and Asofarma, support attending meetings from UCB, Abbie and Celgen. Dr. Isidoro González has received grants or contracts from Grebro Pharma and European Union also payments or honoraria for presentations from Lilly and support for attending meetings from Lilly and Pfizer. Dr. Rosario García have received grants from Abbvie, Lilly, Sanofi and MSD, consulting fees from Biogen, Galápagos, MSD, Sanofi and Johnson&Johnson, payment honoraria for presentations from Pfizer and Abbvie, support for attending meetings from Abbvie, Lly, MSD, Pfizer, Sandoz and UCB and receipt of equipment for her institution from Abbvie. All of them unrelated to the study presented in this manuscript.

Data availability statement:

## Supplementary material

(Supplementary material comprises of data (including text), tables, and figures, and should be referenced in the following format: Supplementary Data S1, Supplementary Table S1, Supplementary Figure S1.)

## Notes

### Author Declarations

The study adhered to ethical principles in biomedical research and the current legislation in Spain, following Good Clinical Practice (ICH Topic E6, 2011), the Declaration of Helsinki, and the requirements of RD 223/2004. The Autonomous Ethics Committee of Galicia (CEIC) approved the study, and all patients provided written informed consent.

