## Supplementary material for "Clinical and Molecular Data to Predict Flares in DMARD optimization in Rheumatoid Arthritis: A Randomised, Controlled, Open-label, Non-inferiority Trial"

**Supplementary material 1.** Optimization protocol

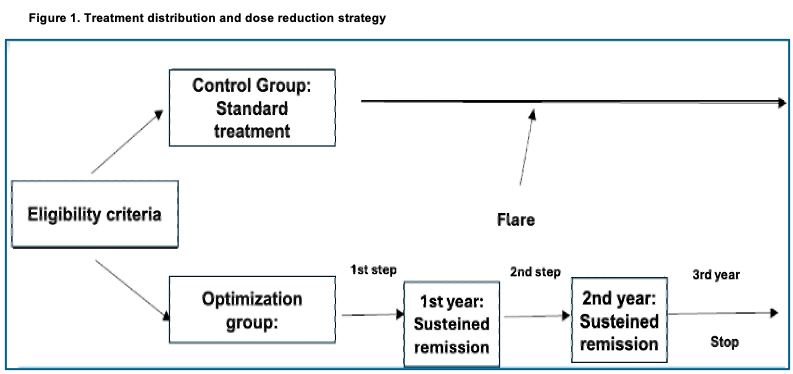

**Supplementary material 2**. List of sequenced genes
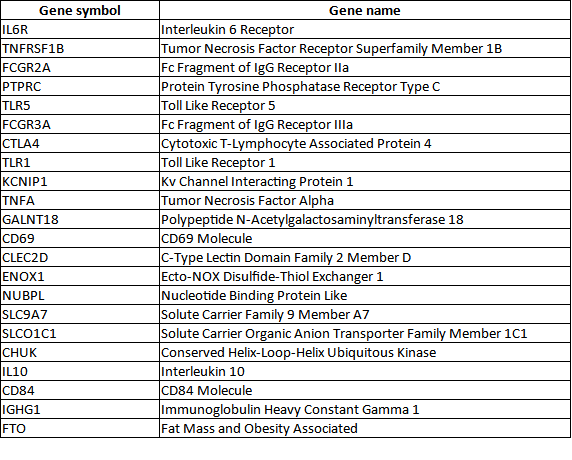

**Supplementary material 3. Targeted-next generation sequencing**

For this study, we used the Ampliseq DNA Panel IAD211967_182 (ThermoFisher Scientific), a NGS-based panel where the individuals exact genotype at all sites is read out across exonic and UTR regions of interest. The panel consisted of a mixture of 22 genes based on extensive curation of peer-reviewed literature and disease research databases. For the sequencing of these 22 genes, the panel consisted of two primer pools (227 primer pairs each) and comprised 454 amplicons. The in-depth sequencing of this gene panel was performed in a subset of 195 genomic DNA (gDNA) samples, of which 96 were samples from patients of the optimization group, and 99 corresponded to samples from patients of the control group.

The preparation of libraries and the subsequent in-depth sequencing of the samples was carried out in the Ion S5XL/Ion Chef platform (ThermoFisher Scientific). A total of 100ng of DNA in a final volume of 15µL were used to construct the barcoded DNA libraries using the Precision ID DL8 Kit (ThermoFisher Scientific) following manufacturer´s recommendations. Up to 8 libraries per run were automatically prepared by the Ion Chef, resulting in a final pool of 700µL at 100pM in one tube. Since the maximum number of barcodes available using this kit is 32, we performed 4 independent runs on the Ion Chef to generate 4 pools of libraries (8 per pool). Finally, each set of (8x4) 32 libraries was pooled into a single tube in a final volume of 25µL and normalized to 70pM, ready for templating and sequencing on the Ion S5XL platform.

Generated libraries were ready for template preparation through clonal amplification on the Ion Chef system following manufacturer´s recommendations. This process concludes with the automatic loading of the samples into the Ion 530 sequencing chip. As a result, approximately 20 million reads per chip were obtained, with an average read depth of 1300X.

**Supplementary material 4.** Analysis of sequencing data

The obtained reads were cleaned and aligned to GrCh37 Human reference sequence using the Torrent Suite 5.18.1 (ThermoFisher Scientific). Variant calling was performed through an adapted pipeline for Ampliseq libraries using a low stringency mode in Torrent Variant Caller plug-in (TVC) (Generic - S5/S5XL (550) - Germ Line - Low Stringency). Each variant was examined based on read depth (≥ 100), Phil´s Read Editor (PHRED) score (≥ 100) and variant frequency (≥ 0.20). The different genomic variant call formats (vcfs) were aggregated using bcftools v1.16 to obtain a multisample vcf containing all the samples in the study. This multisample vcf was filtered to rule out monoallelic variants and variants with a minor allele frequency (MAF) lower than 0.01.

The initial multisample vcf of 195 patients was converted to plink format using plink v1.9 and then performed a logistic regression using either joint flare risk or sustained remission as outcomes. The additive effect of the allelic dose of each variant was included in the model and the results were exported and converted to a bed format using a custom script and subsequently annotated using SnpEff v5.2.

**Supplementary material 5.** Anti-cytokine Autoantibody (ACAA) Analysis:

In this study, a bead-based antigen array (MILLIPLEX® MAP Human Cytokine Autoantibody Magnetic Bead Panel - HCYTAAB-17K, Merck, Darmstadt, Germany), combined with xMAP® technology, was used for the simultaneous detection and quantification of the following 15 anti-cytokine autoantibodies in serum: anti-BAFF, anti-G-CSF, anti-IFNβ, anti-IFNγ, anti-IL-1α, anti-IL-6, anti-IL-8, anti-IL-10, anti-IL-12 (p40), anti-IL-15, anti-IL-17A, anti-IL-17F, anti-IL-18, anti-IL-22, and anti-TNFα.

This multiplex indirect immunoassay enables the detection of antibodies in serum samples using cytokines as antigens immobilized on 15 distinct color-coded magnetic beads. Antigen-antibody interactions are detected using an anti-human immunoglobulin G antibody labeled with the fluorophore phycoerythrin (PE). This assay provides semiquantitative multiplex detection of anti- cytokine IgG antibodies, where the fluorescence signal emitted by phycoerythrin corresponds to the levels of anti-cytokine antibodies in the sample.

The kit also includes four assay control beads (three positive and one negative). The positive controls contain decreasing concentrations of immobilized human IgG: control 1 has the highest concentration, followed by lower concentrations in controls 2 and 3. The negative control comprises beads without immobilized human IgG, serving as a baseline reference. In total, the kit includes 19 different bead regions, each containing the corresponding immobilized cytokine. These were stored in individual vials and prepared according to the manufacturer’s instructions. Briefly, the vials were sonicated for 30 seconds and vortexed for 1 minute. Then, 60 μL from each bead region was transferred into a mixing bottle and resuspended in 6 mL of Assay Buffer (provided in the kit) to create the suspension bead array.

Serum samples were stored at -80°C and thawed at 4°C. Once thawed, they were vortexed and centrifuged at 2,000 g for 1 minute at 4°C to remove suspended particles. Samples were then diluted 1:100 in Assay Buffer and randomized into 96-well plates.

Before sample incubation, the Wash Buffer was diluted 1:10 with deionized water. Each well of a black, clear-bottom 96-well plate was rinsed with 200 μL of this diluted Wash Buffer for 10 minutes at room temperature, followed by removal via decanting. Then, 25 μL of Assay Buffer, 25 μL of the corresponding diluted serum sample, and 25 μL of the suspension bead array (previously vortexed) were added to each well. For background wells, 25 μL of Assay Buffer was added instead of serum. The plate was then sealed, vortexed briefly, and incubated on a plate shaker at 4°C for 16 hours.

After overnight incubation, the plate was washed three times with 200 μL of 1× Wash Buffer using a handheld magnetic plate washer. The plate was allowed to rest on the magnet for 60 seconds to settle the magnetic beads before decanting. Next, 50 μL of the Human Cytokine Autoantibody PE-IgG Conjugate (included in the kit) was added to each well, and the plate was incubated with shaking for 90 minutes at room temperature. Following this, the plate was washed three more times as described above, and 150 μL of Drive Fluid was added to each well. The beads were then resuspended on a plate shaker for 5 minutes and read using a MagPix instrument (Luminex).

The instrument was set to aspirate 100 μL of sample per well. The sample-bead mixture was conveyed via Drive Fluid into the detection chamber, where beads were immobilized into a monolayer using a magnet. The beads were exposed to red and green LEDs: the red LED excited internal dyes used to identify each bead’s color signature, while the green LED (with the RP1 filter) excited the PE fluorophore to measure antibody binding. Filters CL1 and CL2 were used to classify bead identity and eliminate doublets. The instrument was configured to identify a minimum of 50 beads per region.

The fluorescence signals, corresponding to the levels of different anti-cytokine autoantibodies, were reported as median fluorescence intensities (MFI) per bead region, providing a semiquantitative measure of autoantibody levels in each sample. Assay precision was assessed by calculating the coefficient of variation (CV) for replicate samples within each plate. Statistical analyses, including non-parametric tests, Fisher’s exact test, receiver operating characteristic (ROC) curves, and logistic regressions, were performed using SPSS and R. A p-value <0.05 was considered statistically significant.

**Supplementary material 6:** Univariate Cox regression model for flare

| **Demographics characteristics** | | **HR** | | **(IC, 95%)** | **P value** |
| --- | --- | --- | --- | --- | --- |
| Male | | 1·82 | | 0·67-4.95 | 0.238 |
| Age | | 1·04 | | 1·01-1.08 | 0.021 |
| BMI (kg/m2) | | 1·00 | | 0·93-1.08 | 0.994 |
| Smoke (Current or previous smoker) | | 0·59 | | 0·23-1.52 | 0.276 |
| Double positive (ACPA/FR) | | 0·72 | | 0·27-1.94 | 0.523 |
| Positive for anti-citrullinated peptide antibodies (ACPA) | | 0·94 | | 0·39-2.32 | 0.904 |
| Positive rheumatoid factor (RF) | | 0·56 | | 0·24-1.32 | 0.189 |
| Erosions | | 2·25 | | 0·76-6.68 | 0.145 |
| Disease duration, months | | 1·00 | | 1·00-1.00 | 0.408 |
| Remission duration, months | | 1·01 | | 0·99-1.02 | 0.317 |
| **Measures of disease activity** | | | | | |
| Disease Activity Score 28 | | 1·57 | | 0·89-2·77 | 0·119 |
| Disease Activity Score 28 CRP | | 3·57 | | 1·70-7·47 | <0·001 |
| 3v-DAS28 | | 1·37 | | 0·79-2·40 | 0·263 |
| 3v-DAS28-CRP | | 4·98 | | 1·92-12·9 | <0·001 |
| Simplified Disease Activity Index | | 1·14 | | 1·02-1·27 | 0·019 |
| Swollen-joint count 28 | | 1·89 | | 0·25-14·0 | 0·534 |
| Tender-joint count 28 | | 1·59 | | 1·01-2·51 | 0·047 |
| ESR (normal value <20 mm/h) | | 1·00 | | 0·97-1·04 | 0·778 |
| CRP (normal value <1mg/dL) | | 6·01 | | 1·57-23·0 | 0·008 |
| VAS for pain | | 1·03 | | 1·0-1·05 | 0·012 |
| PhGA | | 1·03 | | 1·0-1·06 | 0·036 |
| PGA | | 1·01 | | 1·0-1·03 | 0·134 |
| HAQ-DI | | 1·97 | | 1·14-3·42 | 0·015 |
| **Vitals sing** | |  | |  |  |
| SBP (mm/Hg) | | 1·01 | | 0·99-1·04 | 0·268 |
| Heart Rate (bpm) | | 0·95 | | 0·91-1·0 | 0·046 |
| **Laboratory analysis** | |  | |  |  |
| Hemoglobin | | 1·47 | | 0·95-2·26 | 0·080 |
| Platelets | | 1·00 | | 0·99-1·01 | 0·904 |
| **Molecular variables** | |  | |  |  |
| Anti-TNFi | | 1·00 | | 1·0-1·0 | 0·059 |
| Anti-IFNβ | | 1·00 | | 1·0-1·0 | 0·680 |
| Anti-IFNγ | | 1·00 | | 1·0-1·0 | 0·213 |
| rs11052877 (A/G) | 2.16 | | 1.07-4.33 | | 0.030 |
| rs1560011g (G/A) | 0.40 | | 0.20-0.81 | | 0.010 |
| rs5746065 (C/A) | 0.36 | | 0.16-0.86 | | 0.021 |
| rs6555900 (G/C) | 0.48 | | 0.26-0.90 | | 0.021 |
| rs3093664 (G/A) | 4.14 | | 1.39-12.4 | | 0.011 |
| rs1134543 (G/C) | 3.50 | | 1.07-11.4 | | 0.037 |
| rs9594987 (T/C) | 0.68 | | 0.37-1.28 | | 0.238 |
| ‍ |  | |  | |  |
| BMI= Body-mass index. CRP= C-reactive protein. ERS= Erythrocyte sedimentation rate. 3v-DAS28= Three variables DAS28. PhGA= Physician Global Assessment. PGA= Patient global assessment. HAQ-DI= Health Assessment Questionnaire of Disease Activity | | | | | |

**Supplementary material 7.** Univariate Cox regression analysis of the association between molecular variables and the occurrence of flare in optimization group.

|  | **HR** | **IC95%** | **p** |
| --- | --- | --- | --- |
| **Proteins** | | | |
| Anti-IFNB | 1,000 | 1,000-1,000 | 0,567 |
| Anti-IL22 | 1,000 | 0,999-1,001 | 0,972 |
| Anti-IL12p40 | 1,001 | 0,999-1,002 | 0,273 |
| Anti-IL6 | 1,000 | 1,000-1,000 | 0,749 |
| Anti-IL15 | 1,000 | 0,999-1,001 | 0,869 |
| Anti-IL17A | 1,000 | 1,000-1,000 | 0,992 |
| Anti-IL17F | 1,000 | 1,000-1,000 | 0,619 |
| Anti-GCSF | 1,000 | 0,999-1,001 | 0,863 |
| Anti-TNFa | 1,000 | 1,000-1,000 | 0,112 |
| Anti-IL10 | 1,000 | 0,999-1,000 | 0,391 |
| Anti-BAFF | 1,000 | 1,000-1,000 | 0,560 |
| Anti-IFNG | 1,000 | 1,000-1,001 | 0,001 |
| Anti-IL1a | 1,000 | 1,000-1,000 | 0,859 |
| Anti-IL8 | 1,000 | 0,999-1,001 | 0,420 |
| Anti-IL18 | 1,000 | 0,999-1,001 | 0,541 |
| OD_DSC1 | 0,938 | 0,501-1,759 | 0,843 |
| ngmlDCS1 | 1,000 | 0,999-1,001 | 0,623 |
| **Single nucleotide polimorfism (SNPs)** | | | |
| **rs10108210** | | | |
| AA |  |  |  |
| CA | 1,23 | 0,64-2,36 | 0,53 |
| CC | 1,08 | 0,47-2,52 | 0,851 |
| **rs1071803** | | | |
| CC |  |  |  |
| TC | 0,858 | 0,45-1,62 | 0,637 |
| TT | 0,572 | 0,23-1,43 | 0,234 |
| **rs10919563** | | | |
| AA |  | | |
| GA | 0 | 0-0 | 0 |
| GG |  | | |
| **rs11052877** | | | |
| AA |  | | |
| GA | 0,82 | 0,43-1,56 | 0,546 |
| GG | 2,53 | 1,03-6,17 | 0,0418 |
| **rs11591741** | | | |
| CC |  |  |  |
| CG | 1,19 | 0,51-2,76 | 0,692 |
| GG | 0,865 | 0,35-2,15 | 0,755 |
| **rs1560011** | | | |
| AA |  | | |
| AG | 0,343 | 0,16-0,74 | 0,00672 |
| GG | 0,303 | 0,12-0,74 | 0,00862 |
| **rs1800896** | | | |
| CC |  |  |  |
| CT | 0,798 | 0,27-2,33 | 0,68 |
| TT | 0,918 | 0,31-2,69 | 0,876 |
| **rs3794271** | | | |
| AA |  |  |  |
| GA | 1,36 | 0,72-2,57 | 0,347 |
| GG | 1,02 | 0,4-2,61 | 0,968 |
| **rs4910008** | | | |
| CC |  |  |  |
| CT | 0,757 | 0,4-1,43 | 0,389 |
| TT | 0,523 | 0,21-1,33 | 0,173 |
| **rs6427528** | | | |
| AA |  |  |  |
| GA | 0,000 | 0-Inf | 0,996 |
| GG | 0,000 | 0-Inf | 0,996 |
| **rs703297** | | | |
| CC |  |  |  |
| TC | 1,82 | 0,8-4,17 | 0,156 |
| TT | 1,05 | 0,39-2,84 | 0,917 |
| **rs703505** | | | |
| AA |  |  |  |
| GA | 0,731 | 0,38-1,42 | 0,356 |
| GG | 1,16 | 0,53-2,54 | 0,709 |
| **rs7055107** | | | |
| GG |  | | |
| GT | 0,871 | 0,44-1,73 | 0,693 |
| TT | 0,702 | 0,33-1,5 | 0,361 |
| **rs7195994** | | | |
| AA |  |  |  |
| GA | 1,460 | 0,72-2,95 | 0,294 |
| GG |  |  |  |
| **rs9594987** | | | |
| CC |  |  |  |
| CT | 0,606 | 0,3-1,21 | 0,154 |
| TT | 0,655 | 0,29-1,49 | 0,314 |
| **rs5746065 CA** | | | |
| AA |  |  |  |
| CA | 2,210 | 1,2-4,06 | 0,0108 |
| CC |  |  |  |
| **rs6555900 CG** | | | |
| CC |  |  |  |
| CG | 0,488 | 0,22-1,1 | 0,0851 |
| GG | 0,418 | 0,18-0,96 | 0,0395 |
| **rs3093664 AG** | | | |
| AA |  |  |  |
| AG | 3,21 | 1,35-7,63 | 0,00847 |
| GG |  |  |  |
| **rs115244044AG** | | | |
| AA |  |  |  |
| AG | 0,10 | 0,01-0,77 | 0,0262 |
| GG | 1,47 | 0,2-10,7 | 0,705 |
| **rs2401387 AG** | | | |
| AA |  |  |  |
| AG | 0,418 | 0,2-0,87 | 0,0205 |
| GG | 0,359 | 0,05-2,63 | 0,313 |
| **rs2401388CT** | | | |
| CC |  |  |  |
| CT | 0,418 | 0,2-0,87 | 0,0205 |
| TT | 0,359 | 0,05-2,63 | 0,313 |
| **rs56980650AC** | | | |
| AA |  |  |  |
| AC | 0,418 | 0,2-0,87 | 0,0205 |
| CC | 0,359 | 0,05-2,63 | 0,313 |
| **rs1134543GC** | | | |
| CC |  |  |  |
| GC | 1,17 | 0,15-9,22 | 0,884 |
| GG | 2,79 | 0,38-20,4 | 0,313 |
| **rs6640GT** | | | |
| GG |  |  |  |
| GT | 1,51 | 0,71-3,2 | 0,284 |
| TT | 1,1 | 0,5-2,39 | 0,816 |
| **rs5905551TC** | | | |
| CC |  |  |  |
| TC | 1,27 | 0,66-2,45 | 0,472 |
| TT | 0,592 | 0,26-1,37 | 0,222 |
| **rs6611276CT** | | | |
| CC |  |  |  |
| CT | 1,42 | 0,67-3,01 | 0,36 |
| TT | 1,07 | 0,49-2,33 | 0,869 |
| **rs3208940GA** | | | |
| AA |  |  |  |
| GA | 1,1 | 0,56-2,19 | 0,777 |
| GG | 0,848 | 0,4-1,81 | 0,672 |
| **rs231775AG** | | | |
| AA |  |  |  |
| AG | 0,892 | 0,47-1,67 | 0,722 |
| GG | 1,35 | 0,51-3,59 | 0,546 |
| **rs5743611CG** | | | |
| CC |  |  |  |
| CG | 0,816 | 0,32-2,07 | 0,669 |
| GG | 0,816 | 0,11-5,95 | 0,841 |

**Supplementary material 8** Univariate logistic regression analysis to sustained remission

| **Demographics characteristics** | **OR** | **(IC, 95%)** | **p value** |
| --- | --- | --- | --- |
| Male | 0·53 | 0·16-1·89 | 0·301 |
| Age | 0·95 | 0·90-0·98 | 0·011 |
| BMI (kg/m2) | 1·00 | 0·92-1·09 | 1·000 |
| Smoker | 1·78 | 0·65-5·42 | 0·281 |
| Double positive (ACPA/FR) | 1·38 | 0·43-4·18 | 0·573 |
| Positive for anti-citrullinated peptide antibodies (ACPA | 1·01 | 0·35-2·76 | 0·686 |
| Positive rheumatoid factor (RF) | 1·87 | 0·68-5·03 | 0·217 |
| Erosions | 0·38 | 0·10-1·17 | 0·114 |
| Disease duration, months | 0·99 | 0·99-1·00 | 0·304 |
| Remission duration, months | 1·00 | 0·98-1·04 | 0·536 |
| **Measures of disease activity** | | | |
| Disease Activity Score 28 | 0·56 | 0·27-1·08 | 0·095 |
| Disease Activity Score 28 CRP | 0·19 | 0·06-0·55 | 0·003 |
| 3v-DAS28 | 0·66 | 0·33-1·24 | 0·217 |
| 3v-DAS28-CRP | 0·14 | 0·03-0·49 | 0·003 |
| Simplified Disease Activity Index | 0·85 | 0·72-0·98 | 0·030 |
| Swollen-joint count 28 | 0·58 | 0·05-12·91 | 0·666 |
| Tender-joint count 28 | 0·61 | 0·33-1·11 | 0·103 |
| ESR (mm/hr) | 0·99 | 0·95-1·03 | 0·613 |
| CRP (mg/dL) | 0·12 | 0·02-0·74 | 0·024 |
| VAS for pain | 0·97 | 0·94-0·99 | 0·019 |
| PhGA | 0·96 | 0·92-1·00 | 0·062 |
| PGA | 0·98 | 0·96-1·01 | 0·133 |
| HAQ-DI | 0·38 | 0·17-0·80 | 0·012 |
| **Vital signs** | | | |
| SBP (mm/Hg) | 0·98 | 0·95-1·01 | 0·222 |
| Heart Rate (bpm) | 1·06 | 1·01-1·12 | 0·033 |
| **Laboratory analysis** | | | |
| Hemoglobin (g/dL) | 0·67 | 0·40-1·06 | 0·107 |
| Platelets (mcL) | 1·00 | 0·99-1·00 | 0·932 |
| **Molecular variables** | | | |
| rs11052877 (G/T) | 0.40 | 0.17-0.89 | 0.029 |
| **rs1560011 (G/A)** | 2.91 | 1.30-7.10 | 0.013 |
| **rs5746065 (C/A)** | 3.45 | 1.23-9.80 | 0.018 |
| **rs6555900 (G/C)** | 2.31 | 1.14-4.93 | 0.024 |
| rs3093664 (G/C) | 0.18 | 0.03-0.91 | 0.037 |
| rs2401387 (G/A) | 4.11 | 1.33-18.21 | 0.029 |
| rs1134543 (G/C) | 0.24 | 0.05-0.75 | 0.029 |
| BMI= Body-mass index. CRP= C-reactive protein. ERS= Erythrocyte sedimentation rate. 3v-DAS28= Three variables DAS28. PhGA= Physician Global Assessment. PGA= Patient global assessment. HAQ-DI= Health Assessment Questionnaire of Disease Activity | | | |

**Supplementary material 9:** Univariate logistic regression analysis of the association between molecular variables and the sustained remission in optimization group.

|  | **OR** | **95% CI** | **p** |
| --- | --- | --- | --- |
| **Proteins** | | | |
| Anti-IFNB | 1,00 | 0,99-1,00 | 0,678 |
| Anti-IL22 | 1,00 | 0,99-1,00 | 0,229 |
| Anti-IL12p40 | 0,99 | 0,99-1,00 | 0,659 |
| Anti-IL6 | 1,00 | 0,99-1,00 | 0,173 |
| Anti-IL15 | 1,00 | 1,00-1,01 | 0,215 |
| Anti-IL17A | 1,00 | 0,99-1,00 | 0,846 |
| Anti-IL17F | 1,00 | 0,99-1,00 | 0,246 |
| Anti-GCSF | 1,00 | 0,99-1,00 | 0,236 |
| Anti-TNFa | 1,00 | 1,00-1,00 | 0,056 |
| Anti-IL10 | 1,00 | 1,00-1,00 | 0,100 |
| Anti-BAFF | 1,00 | 0,99-1,00 | 0,487 |
| Anti-IFNG | 0,99 | 0,99-1,00 | 0,254 |
| Anti-IL1a | 1,00 | 0,99-1,00 | 0,878 |
| Anti-IL8 | 1,00 | 0,99-1,00 | 0,856 |
| Anti-IL18 | 1,00 | 0,99-1,00 | 0,252 |
| OD_DSC1 | 1,99 | 0,70-9,50 | 0,277 |
| ngmlDCS1 | 1,00 | 0,99-1,00 | 0,365 |
| **SNPs (Allele additives)** | | | |
| rs1071803t | 1,81 | 0,91-3,91 | 0,106 |
| rs10919563g | 0,77 | 0,21-2,24 | 0,653 |
| rs11052877g | 0,40 | 0,17-0,89 | 0,029 |
| rs11591741g | 1,48 | 0,74-3,03 | 0,269 |
| rs1560011g | 2,91 | 1,30-7,10 | 0,013 |
| rs1800896t | 1,05 | 0,48-2,25 | 0,897 |
| rs3794271g | 0,85 | 0,42-1,73 | 0,647 |
| rs4910008t | 1,14 | 0,57-2,32 | 0,718 |
| rs6427528g | 0,46 | 0,13-1,24 | 0,165 |
| rs703297t | 1,12 | 0,54-2,33 | 0,758 |
| rs703505g | 1,37 | 0,70-2,79 | 0,372 |
| rs7055107t | 0,97 | 0,52-1,80 | 0,921 |
| rs7195994g | 1,04 | 0,27-3,38 | 0,947 |
| rs9594987t | 1,48 | 0,75-3,02 | 0,265 |
| rs5746065 c | 3,45 | 1,23-9,80 | 0,018 |
| rs6555900 g | 2,31 | 1,14-4,93 | 0,024 |
| rs3093664 g | 0,18 | 0,03-0,91 | 0,037 |
| rs115244044g | 3,86 | 0,76-70,87 | 0,197 |
| rs2401387 g | 4,11 | 1,33-18,21 | 0,029 |
| rs2401388t | 4,11 | 1,33-18,21 | 0,029 |
| rs56980650c | 4,11 | 1,33-18,21 | 0,029 |
| rs1134543g | 0,24 | 0,05-0,75 | 0,029 |
| rs6640t | 1,46 | 0,79-2,75 | 0,231 |
| rs5905551t | 1,00 | 0,54-1,86 | 0,992 |
| rs6611276t | 1,50 | 0,81-2,85 | 0,201 |
| rs3208940g | 0,71 | 0,38-1,30 | 0,274 |
| rs231775g | 1,45 | 0,69-3,26 | 0,345 |
| rs5743611g | 1,95 | 0,57-12,22 | 0,365 |

**Supplementary material 10: Adverse events.**

|  | **Standard Group**  **(n=99)** | **Optimization Group**  **(n=96)** |  |
| --- | --- | --- | --- |
| **Adverse Event** | **n (%)** | **N (%)** | **P value** |
| **None** | **88 (**88,89**)** | **86 (**88,66**)** | 0.577 |
| Total | 11 (11.11) | 11 (11.34) |  |
| Any type of infection | 4 (4,04) | 2 (2.06) |  |
| Cancer | 3 (3.03) | 1 (1.03) |  |
| CVE | 1 (1.01) | 3 (3.09) |  |
| Death | 0 (0) | 1 (1.03) |  |
| Other | 3 (3.03) | 4 (4.12) |  |
| Abbreviations: CVE; cardiovascular events | | |  |
